# Cingulate and Frontopolar Cortical Projections to the Cerebellar Vermis Support Prolonged Reaction Time in Identifying Negative Emotional Scenes in Women

**DOI:** 10.1101/2025.01.27.25321160

**Authors:** Hak Kei Wong, Shefali Chaudhary, Yu Chen, Jaime S. Ide, Sheng Zhang, Chiang-Shan R. Li

## Abstract

We previously observed sex differences in the association of individual anxiety and reaction time (RT) during identification of negative emotional scenes in a Hariri task. Prolonged RT in identifying negative (vs. neutral) images represents a behavioral marker of individual anxiety in women but not in men. However, the neural circuit that supports this behavioral observation remains unclear. Here, with a larger sample (64 men and 62 women), we employed whole-brain regression on individual differences in RT during matching negative vs. neutral images or RT (negative – neutral) and evaluated the results at a corrected threshold. Women but not men showed a significant correlation between individual anxiety and RT (negative – neutral), with a slope test confirming the sex difference. In women alone the cerebellar vermis showed activity in positive correlation with RT (negative – neutral). Further, Granger causality mapping (GCM) showed multiple brain regions, including the anterior cingulate cortex/frontopolar cortex (ACC/FPC), that provide inputs to the cerebellar vermis in women. Amongst these regions, only the ACC/FPC cluster showed activity (β) in significant correlation with both STAI State score and RT (negative – neutral) in women. GCM also identified a small cluster in the pons, suggesting that the cortical pontine cerebellar circuit may support prolonged RT during identification of negative emotions. Path analyses further characterized the inter-relationships amongst the neural markers, RT, and anxiety. These findings highlight a behavioral and circuit marker of anxiety state in neurotypical women. Studies with different behavioral paradigms are needed to characterize the behavioral and neural mechanisms of male anxiety.

## 1. INTRODUCTION

### 1.1 Anxiety and negative emotion processing

Dysfunctional negative emotion processing is a hallmark of anxiety state and anxiety disorders, affecting approximately 4% of the global population (Javaid *et al*., 2023). Many studies have combined behavioral paradigms and functional brain imaging to identify these neural processes. For instance, a recent meta-analysis of over 300 patients with anxiety disorders and 300 controls from 20 studies demonstrated elevated activation in core cortical regions of default mode and frontal-parietal networks during negative emotion processing as well as sex and age differences in these regional activities (Wang *et al*., 2024). The authors highlighted the importance of functional disturbances in self-reference and cognitive control as a neural mechanism of anxiety disorders. This along with other meta-analyses and reviews of the literature of fear learning, anticipation of aversive stimuli, and the processing and regulation of emotion (Servaas *et al*., 2013; Xu *et al*., 2019; Akiki *et al*., 2025) elucidates the neural markers of anxiety and suggests potential targets of behavioral treatment in regulating individual anxiety. A more recent work employed a hidden Markov model to identify brain activation states during video watching in adolescents (Camacho *et al*., 2024). With the videos annotated for emotion-specific and nonspecific information and the moment-to-moment probability of being in each brain state, authors showed higher probability of staying in the ventral attention and default mode activation state and lower probability in cingulo-opercular network activation state in individuals with higher levels of generalized anxiety. Thus, imaging studies have implicated the attention, executive control, and default mode circuits in the pathophysiology of dysfunctional anxiety.

### 1.2 Sex differences in anxiety and negative emotion processing

It is well known that clinical manifestations of anxiety vary between men and women, with men often showing externalizing symptoms like anger, aggression or irritability and women internalizing symptoms such as heightened worry and somatic complaints (Altemus, Sarvaiya and Neill Epperson, 2014; Farhane-Medina *et al*., 2022). Furthermore, the rates of lifetime anxiety disorders and subclinical anxiety are approximately twice as high in women as in men (Altemus, Sarvaiya and Neill Epperson, 2014). Other studies suggest critical sex differences in negative emotion processing (Stevens and Hamann, 2012; Li *et al*., 2020a; Chaudhary *et al*., 2023, 2024; Fu *et al*., 2024; Wu *et al*., 2024) and other psychological processes of importance to emotional disorders, such as reward, punishment sensitivity and self-control (Li *et al*., 2006, 2020b; Kann *et al*., 2016; Dhingra *et al*., 2021). Likewise, many studies have reported sex differences in the neural markers of anxiety and depressive disorders (Borst *et al*., 2024; Doucet *et al*., 2024; Hejazi *et al*., 2024; Yang *et al*., 2024; see also Bangasser and Cuarenta, 2021 for a review). For instance, when matching identical facial versus shape stimuli, men as compared with women engaged the occipital-temporal visual cortex and both anterior and posterior cingulate cortex to a greater extent, whereas women relative to men showed higher activation of bilateral middle frontal cortex (Li *et al*., 2020a). These sex differences in regional activities varied widely across behavioral paradigms, so did the neural correlates of anxiety and/or depression as revealed with these behavioral tasks. Although it is not entirely clear what other factors may have accounted for the sex differences, it is clearly important to consider sex as a biological variable in investigating the neural markers of individual anxiety.

### 1.3 A behavioral correlate of anxiety in identification of negative emotion images and the present study

In our previous study, we observed that women with higher anxiety were slower in identifying images of negative relative to neutral emotional scenes in a Hariri task (Chaudhary *et al*., 2024). Thus, individual anxiety state may bias the attention mechanism in women in processing negative emotional stimuli in a way that delays the identification of target images (Prasad, Tarai and Bit, 2024). However, the neural circuits underlying this behavioral marker of anxiety remain unclear.

To address this question, we first identified the regional responses in correlation with prolonged RT during identification of negative vs. neutral emotional images [RT (Neg – Neu)] in a larger sample of women and men. Using the identified brain region as a seed, we performed Granger causality mapping to examine brain regions with significant projections to and/or from the seed and evaluate whether these regional activities were associated with RT (Neg – Neu) and individual anxiety. Finally, we performed mediation and path analyses to characterize the inter-relationship amongst the regional activities, RT (Neg – Neu), and individual anxiety. The goal of the study is to characterize the neural mechanisms of prolonged RT in identifying negative emotional images as a behavioral marker of anxiety.

## 2. METHODS

### 2.1 Participants and assessments

One-hundred-and-twenty-six neurotypical adults (62 women) 19 to 85 years of age volunteered to participate in the study. Participants were recruited from the greater New Haven, Connecticut, area. All were physically healthy with no major medical conditions. Those with current use of prescription medications or with a history of head injury or neurological illness were excluded. Other exclusion criteria included current or history of Axis I disorders according to the Structured Clinical Interview for DSM-IV (First *et al*., 1996). Candidates who reported current use of illicit substances or tested positive for cocaine, methamphetamine, opioids, marijuana, barbiturates, or benzodiazepines were not invited to participate. All participants were assessed with the State-Trait Anxiety Inventory (STAI; Spielberger, 1989). The Human Investigation Committee at Yale School of Medicine approved the study procedures. All participants signed an informed consent prior to the study.

### 2.2 Behavioral task

Participants performed the Hariri task (Chaudhary *et al*., 2024) during functional magnetic resonance imaging (fMRI). This task involved 24 images, divided equally between negative and neutral scenes, presented in a block design. Each trial displayed a target scene at the top, accompanied by two scenes at the bottom, one matching and the other not matching the target. Participants were instructed to match one of the two bottom scenes with the target by pressing either the left or right button using their dominant hand (Fig. 1A). The session began with 10 seconds of dummy scans, followed by a 2-second instruction phase displaying “choose one to match the picture at the top.” This was followed by four blocks: two blocks of negative scenes interspersed with two blocks of neutral scenes, presented in the order: neutral, negative, negative, and neutral. Each block commenced with a 2-second fixation period, followed by six consecutive stimuli, each displayed for 6 seconds without inter-stimulus gaps. The scanning session lasted approximately 152 seconds (∼3 minutes). During the task, participants responded via button presses, allowing for the measurement of accuracy and reaction time (RT). Participants were informed that the stimuli would remain visible long enough to ensure accurate matching and not explicitly instructed to prioritize speed.

**Figure 1.**
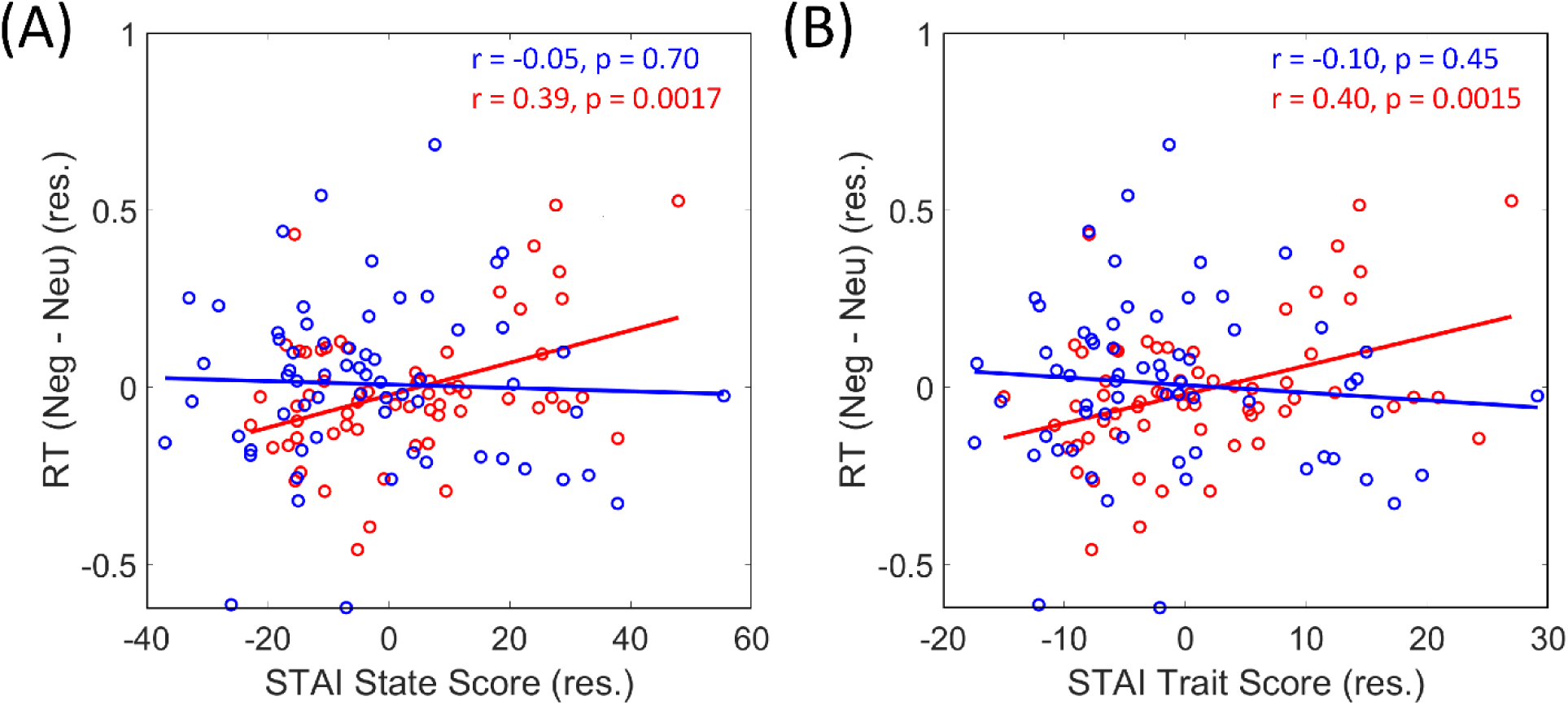
Correlation of RT (Neg – Neu) and STAI (A) State and (B) Trait scores in women (red) and men (blue). Note that, with age as a covariate for the regression, the data points represent the residuals.

### 2.3 Magnetic resonance imaging

Brain images were collected using multiband imaging with a 3-Tesla MR scanner (Siemens Trio, Erlangen, Germany). Conventional T1-weighted spin echo sagittal anatomical images were acquired for slice localization. Anatomical 3D magnetization prepared-rapid gradient echo (MPRAGE) image were next obtained with spin echo imaging in the axial plane parallel to the AC–PC line with TR = 1900 ms, TE = 2.52 ms, bandwidth = 170 Hz/pixel, field of view = 250 × 250 mm, matrix = 256 × 256, 176 slices with slice thickness = 1 mm and no gap. Functional, blood oxygen level-dependent (BOLD) signals were acquired with a single-shot gradient echoplanar imaging (EPI) sequence. Fifty-one axial slices parallel to the AC–PC line covering the whole brain were acquired with TR = 1000 ms, TE = 30 ms, bandwidth = 2290 Hz/pixel, flip angle = 62°, field of view = 210 × 210 mm, matrix = 84 × 84, 51 slices with slice thickness = 2.5 mm and no gap, 392 vol, and multiband acceleration factor = 3. Images from the first ten TRs at the beginning of each scan were discarded to ensure that only BOLD signals in steady-state equilibrium between RF pulsing and relaxation were included in data analyses.

### 2.4 Imaging data analyses

Data were analyzed with Statistical Parametric Mapping (SPM12, Wellcome Department of Imaging Neuroscience, University College London, U.K.), following our published routines (Li et al., 2021, 2020). Briefly, images of each individual subject were first realigned (motion corrected) and corrected for slice timing. A mean functional image volume was constructed for each subject per run from the realigned image volumes. These mean images were co-registered with the high-resolution structural image and segmented for normalization with affine registration followed by nonlinear transformation. The normalization parameters determined for the structure volume were then applied to the corresponding functional image volumes for each subject. The resampled voxel size is 2.5 × 2.5 × 2.5 mm^3^. Finally, the images were smoothed with a Gaussian kernel of 8 mm at Full Width at Half Maximum.

A statistical analytical block design was constructed for each individual subject using a general linear model (GLM) by convolving the canonical hemodynamic response function (HRF) with the boxcar function in SPM, separately for negative and neutral scenes. Realignment parameters in all six dimensions were also entered in the model. The GLM estimated the component of variance that could be explained by each of the regressors. In the first-level analysis, we constructed for each individual subject a contrast of “negative – neutral scene” block to evaluate regions that responded differently to matching these images. The contrast images of the first-level analysis were used for group statistics. In random effects analyses, we performed a one-sample t test to evaluate regional responses to negative vs. neutral images in all participants as well as in men and women separately and a two-sample t test with age as a covariate to evaluate sex differences. We also performed a linear regression of the contrast images against RT (negative – neutral) across all subjects, with age as a covariate, to evaluate neural correlates of the differences in RT for the whole brain. Following current reporting standards, all analyses were evaluated at voxel p<0.001, uncorrected, in combination with cluster p<0.05, FWE corrected, on the basis of Gaussian random field theory (RFT) as implemented in SPM.

### 2.5 Granger causality mapping (GCM)

The methods of GCM was described in detail in our earlier work (Ide and Li, 2011). With the brain region (cerebellar vermis, see Results) identified from whole-brain regression analyses of the correlates of RT (Neg – Neu) as a seed (see Results), we performed GCM (Goebel *et al*., 2003) to show its regional inputs and outputs. Granger causal mapping in MRI is a statistical method used to identify directed connectivity between brain regions. It determines whether the activity (BOLD signals) in one region “causes” changes in another by examining the temporal precedence of signals, based on the concept of Granger causality. Essentially, it assesses whether the activity in one region can predict the activity in another over time (Granger *et al*., 2001; Goebel *et al*., 2003).

Briefly, BOLD time series were entered into a multivariate autoregressive model (Harrison, Penny and Friston, 2003), where the model order indicates how many previous time points are included in the regression. In GCM, we had two time-series *x* and *y,* each representing average BOLD series from the seed and the target regions. To test whether variable *x* Granger causes *y*, we computed the autoregressive model of *y* with and without variable *x*, the *unrestricted* and *restricted* models, respectively. The influence from *x*→*y* can be measured by the fractional *F-value* (Hamilton, 1994) given by the difference between the residual sum of squares (RSS) of the restricted and unrestricted models. If the difference is significantly different from zero, it means that *x* significantly Granger causes *y* (significant connection). We used model order 1 for Granger causality analysis, as suggested by previous simulation studies (Roebroeck, Formisano and Goebel, 2005), to focus on the hemodynamic responses around 1 s and to avoid model overfitting. We called *F-out* the GC influence (connectivity strength) of the seed on an output region and *F-in* the influence of an input region on the seed. Group results of input and output were generated by counting the number of subjects with significant connections, with a binomial test to determine group-level significance.

Following the identification of the input and output regions – regions of interest (ROI) – of the seed, we employed linear regression of the F values (GC strength) and β (Neg – Neu) on STAI State and Trait score as well as RT (Neg – Neu) for each ROI.

## 3. RESULTS

### 3.1 Behavioral correlates of anxiety

Men showed a STAI State (mean ± SD: 30.2 ± 10.2) and Trait (31.9 ± 10.1) score and women showed a STAI State (33.9 ± 10.2) and Trait (35.9 ± 9.5) score. A two-sample t test with age as a covariate showed no differences in the anxiety scores between men and women (State: t = 1.65, p = 0.101; Trait: t = 1.92, p = 0.058). Both men and women were highly accurate in matching both negative (men: 98.7 ± 5.2%; women: 99.6 ± 1.8%) and neutral (men: 98.8 ± 4.2%; 99.5 ± 2.1%) images and showed no differences in Accuracy (Neg-Neu) (men: –0.1 ± 5.0%; women: 0.1 ± 2.3%; t = 0.4, p = 0.725; two-sample t test with age as a covariate). Men showed an RT of 1608 ± 458 and 1598 ± 434 ms and women showed an RT of 1445 ± 360 and 1442 ± 341 ms in matching negative and neutral images, respectively. Both men and women showed quite a bit of variation in individual differences in RT during matching of negative vs. neutral emotional images or RT (Neg – Neu). Men showed a RT (Neg – Neu) of 10 ± 235 (mean ± SD) and women 0 ± 201 ms. A two-sample t test with age as a covariate showed no sex difference (t = –0.46, p = 0.643).

We performed linear regressions of RT (Neg – Neu) on both STAI state and trait scores, with age as a covariate. Across all subjects, RT (Neg – Neu) was not significantly correlated with STAI state (r = 0.131, p = 0.144) or trait score (r = 0.111, p = 0.217). In men alone, RT (Neg – Neu) was not significantly correlated with STAI state (r = –0.050, p = 0.699) or trait (r = –0.097, p = 0.451) score. However, in women, RT (Neg – Neu) was significantly and positively correlated with both STAI state (r = 0.394, p < 0.002) and trait (r = 0.398, p < 0.002) score. A slope test confirmed the sex differences (state: z = –2.53, p = 0.011; trait: z = –2.87, p = 0.0041). **Figure 1** shows the scatter plots and regression lines. Thus, in women but not in men, higher levels of anxiety were associated with prolonged RT in identifying negative vs. neutral images.

### 3.2 Neural correlates of negative emotion processing

We conducted a one-sample t test of “negative – neutral” blocks for all, men, and women. **Figure 2** shows the results evaluated at voxel p<0.001, uncorrected. The clusters that also met cluster p<0.05, FWE corrected are shown in **Supplementary Table S1**.

**Figure 2.**
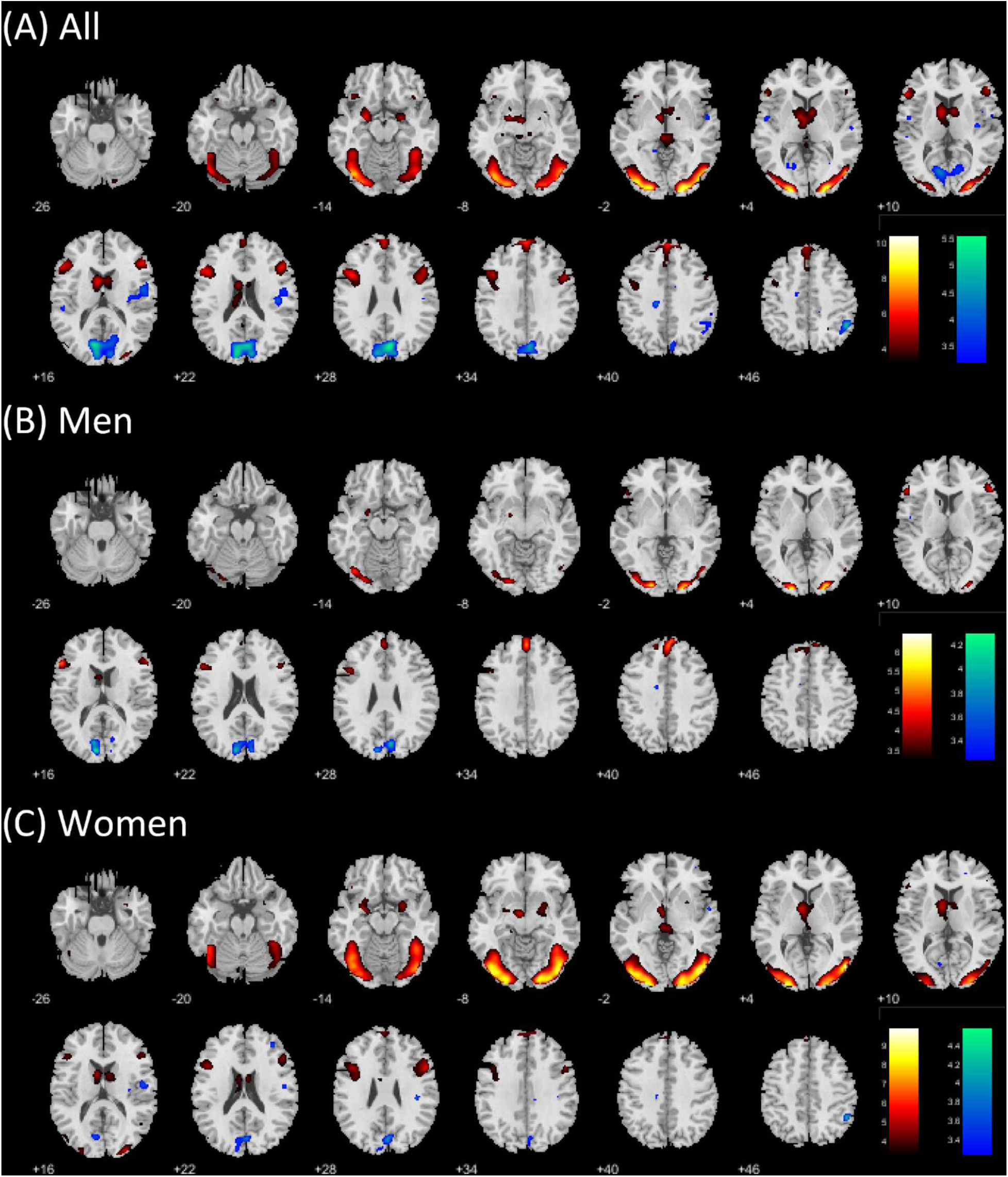
One-sample t test of “negative – neutral” for (A) all; (B) men; and (C) women, evaluated at voxel p<0.001. Warm and cool color each shows negative > neutral and neutral > negative. Color bars show voxel T values. Neurological orientation: right = right.

We conducted a two-sample t test to compare men and women in “negative – neutral” with age as a covariate. Evaluated at the same threshold, no clusters showed significant sex difference.

For the clusters identified from one-sample t test, we derived the β estimates (Neg – Neu) of individual clusters for correlation with RT (Neg – Neu) for all, men, and women with age as a covariate. With a total of 23 clusters, we evaluated the results at a corrected threshold of p=0.05/23=0.0021. None of the Pearson regressions were significant (all p’s > 0.008). The statistics are summarized in **Supplementary Table S2**.

### 3.3 Neural correlates of prolonged RT during negative emotion processing

We employed RT (Neg – Neu) as a regressor in a whole brain regression of the contrast “negative – neutral” with age as a covariate. No clusters met the threshold across all subjects or in men alone. In women, a cluster in the cerebellar vermis (x=8, y=-54, z=-7, voxel Z=4.12, 210 voxels; **Figure 3A**) showed responses in positive correlation with RT (Neg – Neu).

**Figure 3.**
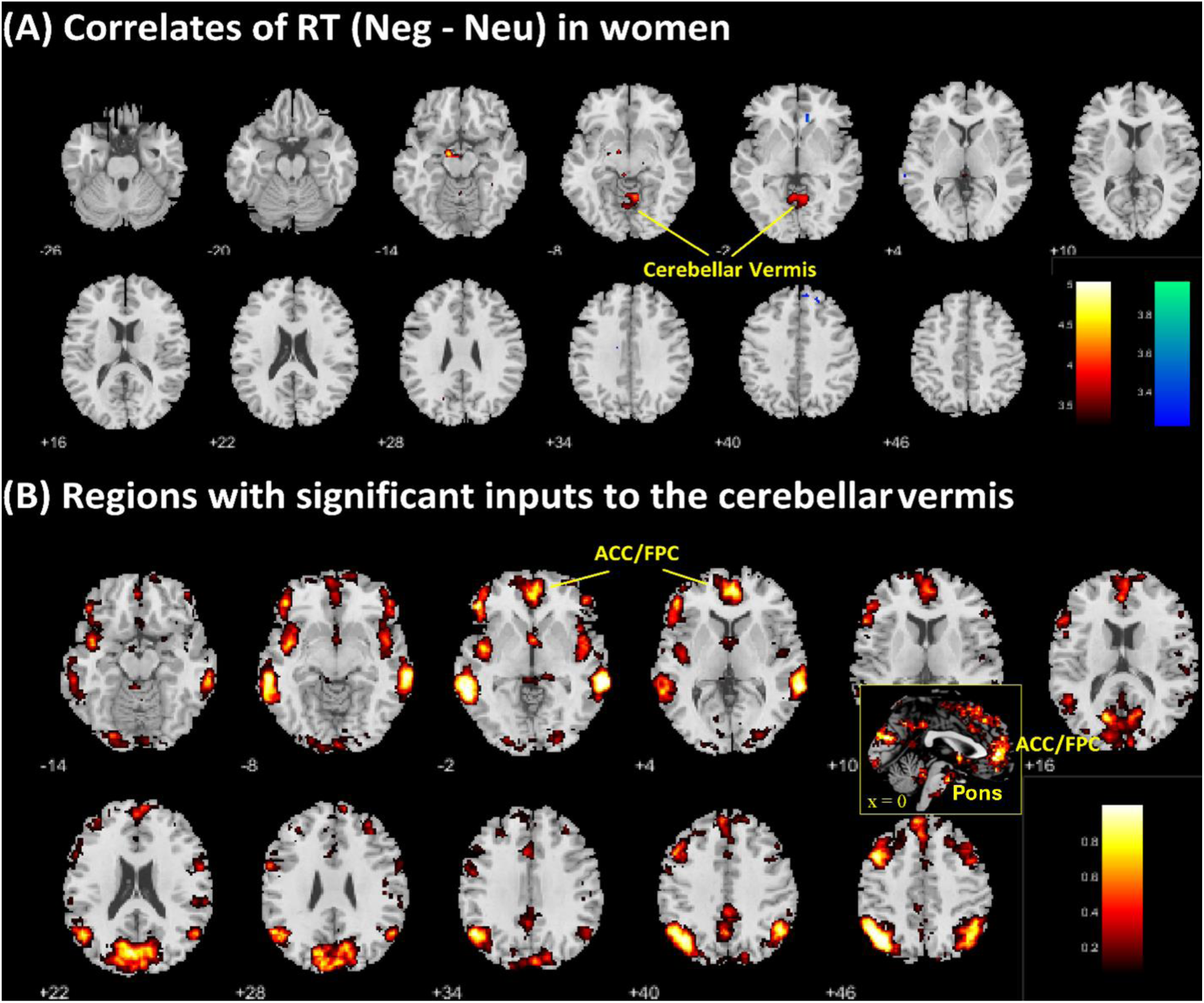
(**A**) Regional correlates of RT (Neg – Neu) in whole brain regression with age as a covariate in women, evaluated at voxel p<0.001. Warm and cool color each shows positive and negative correlation. Color bars show voxel T values. **(B)** Granger causality mapping of brain regions showing significant inputs to the cerebellar vermis as a seed region, evaluated at p<0.05, binomial test. Color bar represents the probability metric of the binomial test.

We computed the β (Neg – Neu) of the cerebellar vermis cluster for all subjects and examined the correlation of the β’s with RT (Neg – Neu) across all subjects and in women and men separately, with age as a covariate. As expected, RT (Neg – Neu) was significantly correlated with the β (Neg – Neu) in women (r=0.47, p<0.001). It also showed a significant correlation for all subjects (r= 0.27, p= 0.0024) but not for men alone (r=0.104, p=0.418). Further, a slope test confirms the sex difference (z=-2.24, p=0.025), suggesting that cerebellar vermis response as a neural correlate of prolonged RT during matching of negative vs. neutral images was sex specific. Note that this analysis did not represent “double-dipping.” As the voxels and clusters were identified with a threshold and those that were identified in women could have just missed the threshold in men, and vice versa. Thus, a post-hoc slope test was needed to confirm sex differences.

We also examined the correlation of the β of the cerebellar vermis with STAI State and Trait scores for all subjects and for men and women separately, with age as a covariate. The results showed that the correlation was significant across all subjects (State: r = 0.22, p = 0.013; Trait: r = 0.27, p = 0.0019) and for women (State: r = 0.37, p = 0.0035; Trait: r = 0.35, p = 0.0056) but not for men (State: r = 0.052, p = 0.69; Trait: r = 0.20, p = 0.11). However, a slope test showed only a trend-level difference between men and women for the correlation with STAI State score (State: z = –1.84, p = 0.066; Trait: z = –0.89, p = 0.37).

Thus, for women alone, the β’s of the cerebellar vermis were significantly correlated with both RT (Neg – Neu) and with STAI state and trait scores. In a mediation analysis, we observed that cerebellar vermis β significantly mediated the correlation between both STAI State/Trait scores and RT (Neg – Neu) in women (**Figure 4**).

**Figure 4.**
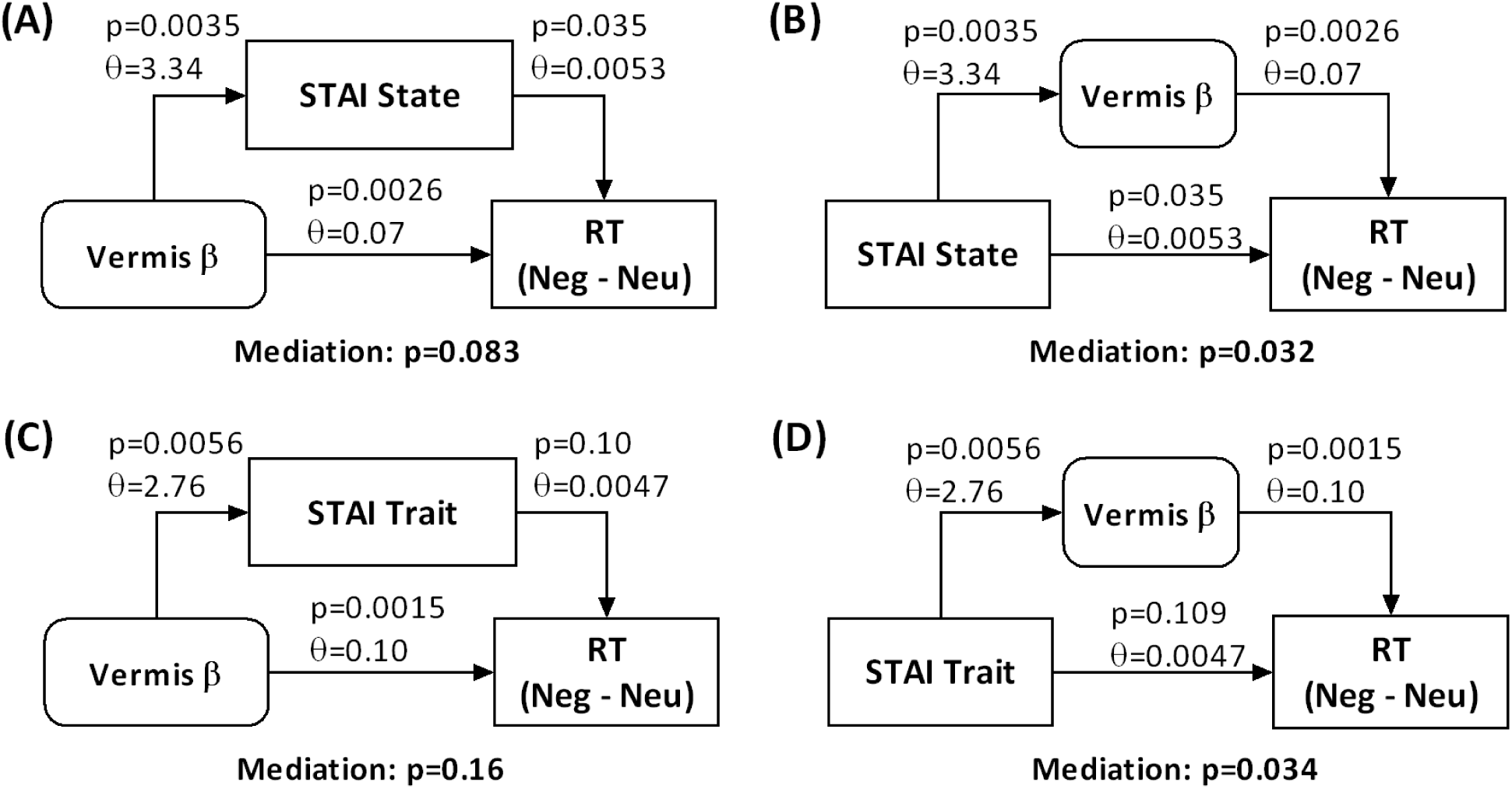
Mediation analyses of the inter-relationship between cerebellar vermis (Vermis) β’s, STAI State (A, B) or Trait (C, D) score, and RT (Neg – Neu) in women.

### 3.4 GCM of cerebellar vermis connectivity during identification of negative emotion in women

We performed Granger causality mapping (GCM) with the cerebellar vermis as the seed region in women. Evaluated with a p<0.05, binomial test (see Methods) and a cluster size of 10 voxels, the results showed a multitude of brain regions with significant inputs to the cerebellar vermis (**Figure 3B**, **Supplementary Table S3**). Evaluated at the same threshold, no clusters were identified as outputs of the cerebellar vermis.

These input regions comprise the saliency circuit and include the anterior cingulate cortex/frontopolar cortex (ACC/FPC), bilateral middle frontal gyri, angular gyri, temporo-parietal junction, and insula. We extracted β (Neg – Neu) of the individual clusters for correlation with RT (Neg – Neu) and with STAI State and Trait scores in women and, for comparison, in men. Of these correlations, only the ACC/FPC cluster showed a significant correlation with RT (Neg – Neu) (r = –0.28, p = 0.027) and STAI State (r = –0.34, p < 0.008) but not Trait (r = –0.18, p = 0.16) score, with age as a covariate, in women. The same regressions were not significant in men (all p’s > 0.49), and slope tests confirmed sex differences in the correlation with STAI State (z = 2.43, p = 0.015). Thus, the ACC/FPC shows regional response to negative vs. neural images in association with individual anxiety and RT (Neg – Neu) and provides inputs to the cerebellar vermis where the response to the same was also significantly associated with both RT (Neg – Neu) and individual anxiety in women.

### 3.5 Path analyses of regional activities, RT (Neg – Neu), and anxiety

The preceding analyses identified cerebellar vermis and ACC/FPC with activities (β’s) in correlation with both individual STAI State score and RT (Neg – Neu). We performed a path analysis to evaluate the inter-relationships amongst the β’s of the cerebellar vermis and ACC/FPC, STAI State score, and RT (Neg – Neu), with age as a covariate, in women. Building on the mediation results and GCM finding of cerebellar vermis receiving inputs from ACC/FPC, the analysis demonstrated optimal fit for two models, where ACC/FPC β’s had an indirect effect on RT (Neg – Neu) via cerebellar vermis β’s and STAI State score had a direct effect or, via ACC/FPC and cerebellar vermis β’s or cerebellar vermis β’s alone, an indirect effect on RT (Neg – Neu), as shown in **Figure 5**. The results of other path models were shown in **Supplementary Figure S1** and all path statistics are shown in **Supplementary Table S4**. Thus, the models suggest that, during a state of anxiety, the vermis may receive reduced input from the ACC/FPC and, as a results, higher activation of the cerebellar circuit to support prolonged RT during the processing of negative versus neutral stimuli.

**Figure 5.**
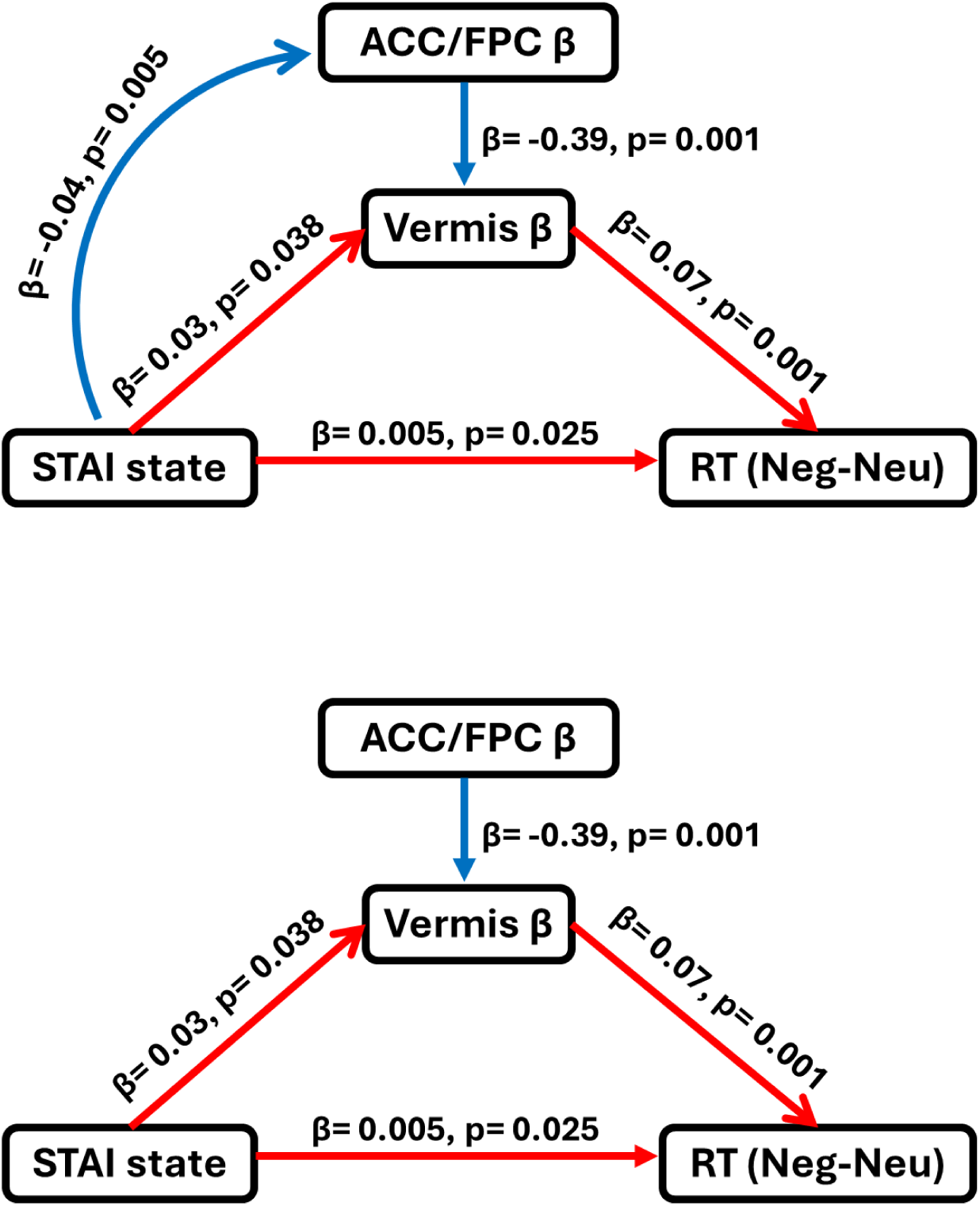
Optimal-fit path models illustrating the interrelationships among the β’s of the cerebellar vermis and ACC/FPC, STAI State score, and RT (Neg – Neu), with age as a covariate, in women. Positive and negative coefficients are represented by paths in ‘red’ and ‘blue,’ respectively. Path statistics are presented as coefficients (β) and p-values.

## 4. Discussion

We aimed to characterize the neural circuits that support individual anxiety and prolonged RT in identifying negative vs. neutral images [RT (Neg – Neu)] in women. With whole-brain regression on RT (Neg – Neu), we identified a cluster in the cerebellar vermis with β (Neg – Neu) in positive correlation with both RT (Neg –Neu) and individual anxiety. Further, Granger causality mapping (GCM) demonstrated a cluster in the anterior cingulate and frontopolar cortex (ACC/FPC) with β (Neg – Neu) in negative correlation with both RT (Neg –Neu) and individual anxiety. GCM also showed a small cluster in the pontine nuclei, suggesting that the circuit ACC → pontine nuclei → cerebellar vermis (and potentially → deep cerebellar nuclei → spinal cord) may support prolonged RT in identifying negative emotional images as a behavioral marker of individual anxiety in women. We discuss the main findings in the below.

### 4.1 Cingulate/frontopolar cortical cerebellar projection and emotional processing

Prefrontal cortex projects to the pontine nuclei, which relay signals to the cerebellum via the middle cerebellar peduncle. Through the superior cerebellar peduncle, the cerebellar cortex projects to the deep cerebellar nuclei, which then project to the thalamus and in turn a wide swath of prefrontal cortical regions. This topographically organized cortico-pontine-cerebellar-thalamic-cortical circuit supports a multitude of cognitive and emotional motor processes (Middleton and Strick, 2001), including motor planning (Zhu *et al*., 2023). In particular, beyond cerebellum’s well-known function in motor control, cerebellar connections with the limbic circuits regulate the experience and expression of emotion as well as physiological responses associated with the emotional experiences (Blatt, Oblak and Schmahmann, 2013).

Amongst these cortical regions, the anterior cingulate cortex (ACC) and frontopolar cortex (FPC) have been widely investigated for their roles in emotion processing, including goal-directed behavior in affect-laden contexts (Etkin, Egner and Kalisch, 2011; Rolls, 2019; Ziaei *et al*., 2019). For instance, the ACC is involved in motor inhibition during a valenced emotional state in a go/nogo task (Albert *et al*., 2012). The FPC is involved in update of emotional memory (Sakaki, Niki and Mather, 2011), and failure in recruiting the FPC to control emotional behavior may account for individual anxiety (Bramson *et al*., 2023). The FPC is involved in representing the congruency of emotion-action goals – faster go RT and higher nogo accuracy each during an appetitive and aversive emotional state – suggesting a role of the FPC in integrating emotional states and goal representations for motor action (Lapate *et al*., 2022). In clinical studies, ACC dysfunction is widely implicated in pathophysiology of dysregulated emotional states, including anxiety and depression (García-Cabezas and Barbas, 2017; Xiao and Zhang, 2018). Greater activation of the FPC during reappraisal of negative emotions during the course of psychotherapy was associated with symptom improvement in people with anxiety disorders (Fonzo *et al*., 2017).

Thus, the current findings of lower ACC/FPC response to matching of negative vs. neutral emotional images in association with higher levels of individual anxiety is consistent with this literature. The current findings extend this literature by characterizing a cortical-cerebellar circuit mechanism of prolonged RT in matching negative vs. neutral emotional images, a behavioral marker of individual anxiety.

### 4.2 Sex differences in the circuit mechanism of prolonged RT in matching negative vs. neutral emotions

Notably, we identified this circuit mechanism that supports prolonged RT in matching negative vs. neutral emotional images as a behavioral marker of individual anxiety only in women. The “null” finding in men seems unlikely to reflect an issue of statistical power, as an approximately equal number of men (n=64) and women (n=62) participated in the study. Men and women also showed similar extent of individual variation in RT (Neg – Neu) to allow regression analysis of the sample. This “lack of finding” in men has also been reported in earlier studies of emotion tasks. For instance, in an MR imaging study of emotion regulation with cognitive reappraisal as a strategy, females showed different levels of self-rated emotional intensity and amygdala activity for negative versus positive emotions, while males did not. Further, females showed greater overall prefrontal cortical but similar levels of amygdala activity compared to males (Min *et al*., 2023). Conversely, other studies reported null findings in women. For instance, men but not women showed a significant, positive correlation between ventral striatal loss reactivity in a guessing task and externalizing traits, with the sex difference confirmed by a slope test (Li *et al*., 2023). A majority of studies reported significant, sex-specific imaging findings in both men and women, including, for instance, studies of facial emotion processing and correlates of anger and fear traits (Li *et al*., 2020a) as well as reward response in the monetary incentive delay task and correlates of reward and punishment sensitivity (Dhingra *et al*., 2021). With these considerations, it would seem most appropriate to conclude that RT (Neg – Neu) or the Hariri task or likely other behavioral paradigms that simply involve exposure to negative emotions may not be ideal for elucidation of the mechanisms of emotion processing or the identification of a behavioral or neural marker of individual anxiety in men.

Men and women showed differences in physiological responses to emotional exposure while watching videos, with women reporting higher levels of arousal and stronger avoidance motivation than men when exposed to negative emotions (Deng *et al*., 2016). Consistently, in an event-related potential (ERP) study, women showed greater ERP to unpleasant as compared with pleasant slides whereas men had greater ERP to pleasant vs. neutral slides (Bianchin and Angrilli, 2012). Further, women relative to men perceived all slides as less pleasant and reported greater arousal to unpleasant condition. In an MR imaging study that required participants to reappraise negative emotions, men relative to women showed less elevation in prefrontal cortical activities that are associated with reappraisal but greater decrement in amygdala activities, which are associated with emotional responding (McRae *et al*., 2008). The latter findings suggest that men may expend less effort in employing cognitive regulation but somehow were able to mitigate their emotional responses. The sex differences in subjective, physiological, perceptual responses to negative emotions were also reported in many other studies (Gard and Kring, 2007; Gohier *et al*., 2013; Poláčková Šolcová and Lačev, 2017). However, the exact behavioral and neurobiological underpinnings of the sex differences remain unclear. Candidate mechanisms for this male “advantage” include expressive suppression that can perhaps be automatized (Cai *et al*., 2016). To reveal how men process negative emotions would likely require behavioral paradigms that involve active processing beyond automatized suppression of emotion, such as those of working memory and complex stimulus-response association learning. Studies are needed to test this hypothesis.

### 4.3 Limitations of the study

We consider several limitations in view of the current findings. First, our sample comprised neurotypical individuals and candidates with clinical anxiety or depression were not recruited for the study. Thus, the current findings should be considered as specific to this population. Second, our sample ranged widely in age. Although we considered age as a covariate in all analyses and one could argue that the findings obtained of the sample may generalize to this age range, how age impacts the current findings remained to be clarified. Finally, as discussed earlier, the “null” findings in men should be interpreted as specific to the behavioral paradigm. More studies that require active engagement in negative emotions are needed to investigate the behavioral and neural markers of anxiety in men.

### 4.4 Conclusions

To conclude, with a direct regression on the difference in RT in matching negative vs. neutral emotional images or RT (Neg – Neu), we identified cerebellar vermis response as a neural correlate of both the RT marker and individual anxiety. As revealed by Granger causality mapping, the anterior cingulate/frontopolar cortex also shows response in correlation with both RT (Neg – Neu) and anxiety state. Together, the findings support a cortico-cerebellar circuit in supporting prolong RT as a behavioral marker of individual anxiety in women.

## Data Availability

All data produced in the present study are available upon reasonable request to the authors

## Acknowledgements

The current study was supported by NIH grants R21AG067024 (Li) and R01AG072893 (Li). The NIH is otherwise not responsible for the design of the study or data analyses and interpretation or in the decision to publish these findings.

