## Supplement for "Cingulate and Frontopolar Cortical Projections to the Cerebellar Vermis Support Prolonged Reaction Time in Identifying Negative Emotional Scenes in Women"

**Supplementary Table S1.** Clusters identified of one-sample t test of "negative – neutral" (Figure 2) that met cluster p<0.05, FWE corrected.

| volume (voxels) | peak voxel (Z) | MNI coordinates (mm) x | y | z | side | identified brain region |
|---|---|---|---|---|---|---|
| *Positive (all)* | | | | | | |
| 1,331 | Inf | -20 | -96 | 1 | L | Occipital cortex |
| 1,503 | Inf | 23 | -96 | 1 | R | Occipital cortex |
| 981 | 6.29 | 5 | 57 | 38 | L/R | dmPFC |
| 710 | 5.76 | -43 | 14 | 28 | L | Inferior frontal gyrus |
| 442 | 5.57 | 53 | 32 | 13 | R | Inferior frontal gyrus |
| 1,132 | 5.34 | -23 | -1 | -15 | L/R | Amygdala, caudate |
| 155 | 4.43 | -3 | -29 | -2 | L/R | |
| *Negative (all)* | | | | | | |
| 1,176 | 5.23 | 5 | -76 | 26 | L/R | Cuneus |
| 163 | 4.92 | 50 | -51 | 48 | R | Inferior parietal gyrus |
| 241 | 4.05 | 43 | -19 | 23 | R | Postcentral gyrus |
| *Positive (men)* | | | | | | |
| 455 | 5.62 | -18 | -96 | 1 | L | Occipital cortex |
| 250 | 5.39 | 23 | -96 | 1 | R | Occipital cortex |
| 389 | 4.70 | 0 | 49 | 36 | L/R | dmPFC |
| 228 | 4.67 | -55 | 27 | 18 | L | Inferior frontal gyrus |
| *Negative (men)* | | | | | | |
| 344 | 4.01 | -10 | -86 | 18 | L/R | Cuneus |
| *Positive (women)* | | | | | | |
| 1,865 | 7.57 | 50 | -76 | 3 | R | Occipital cortex |
| 1,660 | 7.15 | -23 | -99 | 3 | L | Occipital cortex |
| 369 | 5.55 | 8 | 59 | 33 | L/R | dmPFC |
| 763 | 4.98 | -5 | -9 | -7 | L | Caudate |
| 259 | 4.73 | 53 | 14 | 28 | R | Inferior frontal gyrus |
| 303 | 4.52 | -45 | 14 | 23 | L | Inferior frontal gyrus |
| 181 | 4.29 | 25 | 4 | -12 | R | Amygdala |

| | | | | | | |
|---|---|---|---|---|---|---|
| *Negative (women)* | | | | | | |
| 197 | 3.83 | 5 | -76 | 26 | L/R | Cuneus |

Note: R/L: right/left; dmPFC: dorsomedial prefrontal cortex

**Supplementary Table S2**. The results of linear regression of the β's of "negative – neutral" on RT (Neg – Neu) for all clusters identified from one-sample t test of all, men, and women.

| Individual clusters identified | | all | | Men | | Women | |
|---|---|---|---|---|---|---|---|
| **From** | | r | p | r | p | r | p |
| **All:** | 'Positive_-3_-29_-2_roi.mat' | 0.21 | 0.021 | 0.11 | 0.367 | 0.33 | 0.009 |
| | 'Positive_5_57_38_roi.mat' | -0.08 | 0.385 | -0.03 | 0.825 | -0.14 | 0.279 |
| | 'Positive_-20_-96_1_roi.mat' | 0.17 | 0.061 | 0.14 | 0.255 | 0.22 | 0.092 |
| | 'Positive_-23_-1_-15_roi.mat' | 0.10 | 0.286 | -0.01 | 0.923 | 0.26 | 0.043 |
| | 'Positive_23_-96_1_roi.mat' | 0.19 | 0.032 | 0.19 | 0.133 | 0.21 | 0.099 |
| | 'Positive_-43_14_28_roi.mat' | 0.08 | 0.398 | 0.08 | 0.546 | 0.08 | 0.552 |
| | 'Positive_53_32_13_roi.mat' | 0.15 | 0.091 | 0.14 | 0.255 | 0.16 | 0.202 |
| | 'Negative_5_-76_26_roi.mat' | 0.01 | 0.942 | 0.04 | 0.752 | -0.03 | 0.810 |
| | 'Negative_43_-19_23_roi.mat' | 0.00 | 0.977 | 0.08 | 0.542 | -0.12 | 0.342 |
| | 'Negative_50_-51_48_roi.mat' | -0.02 | 0.825 | 0.01 | 0.915 | -0.07 | 0.582 |
| **Men:** | 'Positive_0_49_36_roi.mat' | -0.16 | 0.073 | -0.07 | 0.599 | -0.27 | 0.035 |
| | 'Positive_-18_-96_1_roi.mat' | 0.16 | 0.068 | 0.15 | 0.237 | 0.19 | 0.133 |
| | 'Positive_23_-96_1_roi.mat' | 0.18 | 0.038 | 0.14 | 0.253 | 0.26 | 0.042 |
| | 'Positive_-55_27_18_roi.mat' | 0.06 | 0.521 | 0.06 | 0.639 | 0.06 | 0.665 |
| | 'Negative_-10_-86_18_roi.mat' | -0.03 | 0.724 | 0.00 | 0.997 | -0.07 | 0.604 |
| **Women:** | 'Positive_-5_-9_-7_roi.mat' | 0.12 | 0.185 | 0.01 | 0.936 | 0.30 | 0.020 |
| | 'Positive_8_59_33_roi.mat' | -0.05 | 0.602 | -0.03 | 0.814 | -0.07 | 0.614 |
| | 'Positive_-23_-99_3_roi.mat' | 0.18 | 0.048 | 0.15 | 0.248 | 0.24 | 0.061 |
| | 'Positive_25_4_-12_roi.mat' | 0.06 | 0.529 | 0.04 | 0.759 | 0.10 | 0.432 |
| | 'Positive_-45_14_23_roi.mat' | 0.13 | 0.155 | 0.12 | 0.334 | 0.14 | 0.278 |
| | 'Positive_50_-76_3_roi.mat' | 0.20 | 0.028 | 0.19 | 0.124 | 0.22 | 0.087 |
| | 'Positive_53_14_28_roi.mat' | 0.16 | 0.072 | 0.14 | 0.254 | 0.19 | 0.139 |
| | 'Negative_5_-76_26_roi.mat' | -0.04 | 0.631 | 0.00 | 0.972 | -0.10 | 0.440 |

**Supplementary Table S3.** Brain regions with significant inputs to the cerebellar vermis in Granger causality mapping.

| volume ($mm^3$) | MNI coordinates (mm) | | | identified brain region |
|---|---|---|---|---|
| | x | y | z | |
| 27,703 | 0 | -79 | 22 | Cuneus |
| 281 | -12 | 44 | 49 | Superior Frontal Gyrus |
| 500 | -19 | 63 | 6 | Superior Frontal Gyrus |
| 203 | -2 | 20 | 31 | ACC |
| 14,375 | -32 | 22 | 49 | Middle Frontal Gyrus |
| 18,172 | -45 | -60 | 37 | Angular Gyrus |
| 14,625 | -46 | 24 | 0 | Inferior Frontal Gyrus/Insula |
| 1,484 | -4 | -97 | -8 | Calcarine |
| 13,609 | -60 | -39 | -5 | Middle Temporal Gyrus |
| 687 | 17 | 43 | 47 | Superior Frontal Gyrus |
| 156 | 15 | 2 | 73 | Superior Frontal Gyrus |
| 484 | 19 | -92 | 1 | Calcarine |
| 8,984 | 1 | 49 | 0 | mPFC |
| 312 | 20 | 64 | 15 | Superior Frontal Gyrus |
| 1,000 | 32 | 41 | 29 | Middle Frontal Gyrus |
| 4,922 | 36 | 20 | 48 | Middle Frontal Gyrus |
| 203 | 41 | 1 | 57 | Middle Frontal Gyrus |
| 13,750 | 45 | -57 | 42 | Angular Gyrus |
| 6,844 | 47 | 20 | -5 | Insula |
| 281 | 54 | 28 | 11 | Inferior Frontal Gyrus |
| 437 | 58 | 3 | -22 | Middle Temporal Gyrus |
| 10,875 | 63 | -38 | -5 | Middle Temporal Gyrus |

### Model1

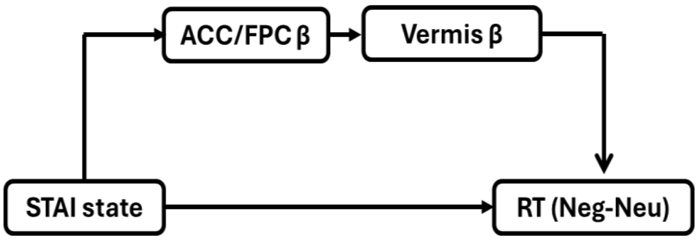


### Model2

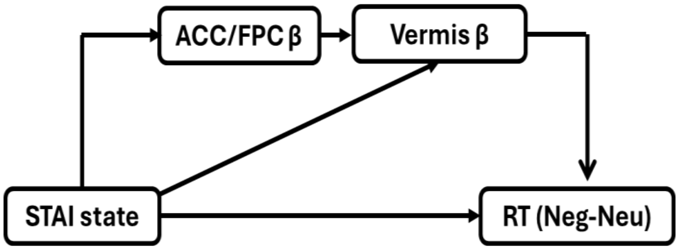


**Model3**

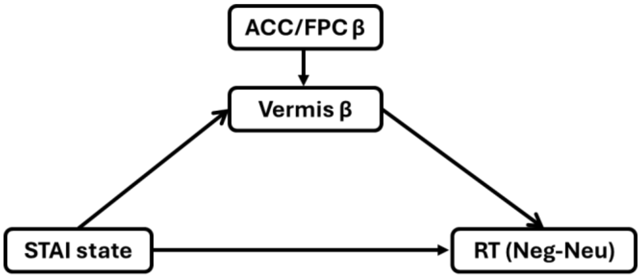


**Model4**

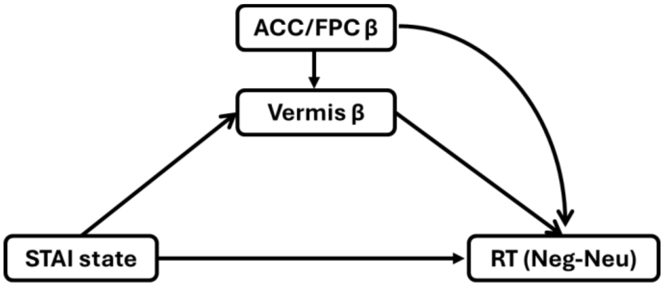


**Model5**

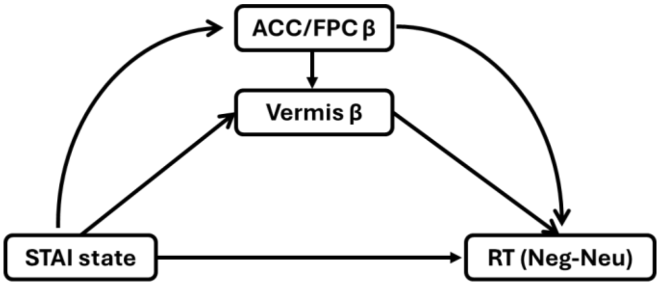


**Supplementary Figure S1:** All tested path models illustrating the interrelationships among the β's of the cerebellar vermis and ACC/FPC, STAI State score, and RT (Neg – Neu), with age as a covariate, in women. Path statistics in Supplementary Table S4.

**Supplementary Table S4.** Statistics of path models

| | Model 1 | Model 2* | Model 3* | Model 4# | Model 5# |
|---|---|---|---|---|---|
| ***Likelihood ratio*** | | | | | |
| $\chi^2$ | 4.241 | 0.062 | 0.062 | 0.000 | 0.000 |
| **p >** $\chi^2$ | 0.120 | 0.803 | 0.803 | - | - |
| ***Population error*** | | | | | |
| **RMSEA** | 0.136 | 0.000 | 0.000 | 0.000 | 0.000 |
| **90% CI, lower bound** | 0.000 | 0.000 | 0.000 | 0.000 | 0.000 |
| **90% CI, upper bound** | 0.318 | 0.214 | 0.214 | 0.000 | 0.000 |
| ***Baseline comparison*** | | | | | |
| **CFI** | 0.949 | 1.000 | 1.000 | 1.000 | 1.000 |
| ***Size of residuals*** | | | | | |
| **SRMR** | 0.056 | 0.006 | 0.006 | 0.000 | 0.000 |
| ***Information criteria*** | | | | | |
| **AIC** | 1305.596 | 1303.417 | 1295.417 | 1297.355 | 1305.355 |
| **BIC** | 1333.249 | 1333.197 | 1316.688 | 1320.753 | 1337.262 |

Note: *Models with optimal fit. #Saturated models are included to demonstrate the maximum possible fit but not for interpretation. A good model fit is typically assessed with indices including $\chi^2$ at p>0.05, root mean square error of approximation (RMSEA)≤0.08, standardized root mean squared residual (SRMR)≤0.08, and comparative fit index (CFI)≥0.90 (Chaudhary *et al.*, 2022).
